# Hope is No Plan: Uncovering Actively Missing Transition-Aged Youth with Congenital Heart Disease

**DOI:** 10.1101/2021.09.21.21263921

**Authors:** Judson A Moore, Shreya S Sheth, Wilson W Lam, Alexander J Alexander, John C Shabosky, Andre Espaillat, Donna K Lovick, Nicole S Broussard, Karla J Dyer, Keila N Lopez

## Abstract

**Background and Objectives:** Studies describing gaps in care for youth with congenital heart disease (CHD), focus on those who have returned to care, but rarely those actively missing from care. Our objective was to determine barriers for young adults with CHD actively missing from cardiac care and to re-engage them in care.

**Methods:** Retrospective single-center cohort study of cardiology clinic patients ages 15-21 years with CHD between 2012-2019 for patients actively missing from care (≥12 months beyond requested clinic follow-up). We conducted prospective interviews, offered clinic scheduling information and recorded cardiac follow-up. Data was analyzed using descriptive statistics, univariable, and multivariable logistic regression.

**Results:** Of 1053 CHD patients, 33% (n=349) were actively missing. Of those missing, 58% were male and median age was 17 yrs (IQR 16-19). Forty-six percent were Non-Hispanic White, 33% Hispanic, and 9% Black. Moderately complex CHD was in 71%, and 62% had private insurance. Patients with simple CHD, older age at last encounter (18-21), and scheduled follow-up >12 months from last encounter were more likely to be actively missing. Interviews were completed by 125 patients/parents (36%). Lack of cardiac care was reported in 52%, and common barriers included: insurance (33%), appointment scheduling (26%), and unknown ACHD center care (15%). Roughly half (55%) accepted appointment information, yet only 3% successfully returned.

**Conclusions:** Many patients require assistance beyond CHD knowledge to maintain and re-engage in care. Future interventions should include scheduling assistance, focused insurance maintenance, understanding where to obtain ACHD care, and educating on need for lifelong care.

**Article Summary:** There is a growing population of youth with congenital heart disease leaving cardiology care; we investigate patients actively missing from care and target their re-engagement.

**What’s Known on This Subject?:** Maintaining cardiac care is a problem for many patients with congenital heart disease, especially teens and young adults, and characteristics of both patients and their healthcare environment contribute to the maintenance and ability to continue appropriate cardiac care.

**What This Study Adds:** This study uniquely investigates patients with congenital heart disease that are actively missing from care, and investigates if a simple intervention of contacting them while they are missing to re-engage them in care impacts their re-entry into cardiac care.

**Contributors’ Statement Page:** Judson Moore conceptualized and designed the study, designed data collection instruments, collected data, drafted the initial manuscript, and completed necessary revisions to final manuscript.

Keila Lopez conceptualized and designed the study, designed data collection instruments, critically reviewed data, revised manuscript drafts, and provided content expertise.

Shreya Sheth conceptualized and designed the study, critically reviewed data, and revised the manuscript.

Wilson Lam contributed to study design, critically reviewed data for intellectual content, and revised the manuscript.

Alexander Alexander, Andre Espaillat, and John Shabosky designed data collection instruments, collected data, and reviewed/revised the manuscript.

Karla Dyer performed data extraction, data cleaning, and reviewed/revised the manuscript.

Nicole Broussard and Donna Lovick provided expert knowledge for interpretation of data and reviewed/revised the manuscript.

All authors approved the final manuscript as submitted and agree to be accountable for all aspects of the work.

## INTRODUCTION

Assuring pediatric patients with chronic diseases maintain medical care to adulthood is a challenge spanning across specialties. Both morbidity and mortality are amplified when follow-up obstacles fail to be addressed^1-5^. This is especially true for patients with chronic conditions that require regular medical encounters over the course of their lifetime, such as those with congenital heart disease (CHD)^1,6-8^. Maintaining care is an even greater issue during adolescence and young adulthood when time, financial, and social constraints are prevalent^1,8,9^. Gaps in care affect up to 60% of CHD patients, and only 10% of adults with CHD are in appropriate care^10^. Individuals with CHD who have fallen out of care may only re-engage in care once symptomatic, potentially allowing for irreversible damage to have already ensued^8^. Previously established barriers to maintaining CHD care have included male sex, less severe disease, and lack of symptoms^8,11^.

The United States has the highest rates of discontinuity of care when compared to Canada and several European countries, with racial/ethnic minorities documented as having worse outcomes with poorer continuity of care and less transition and transfer to adult providers.^9,12^ Other barriers to continuity and transitions of care are the limited access of adult congenital heart disease (ACHD) accredited institutions (institutions that have all the appropriate resources and medical providers to take care of ACHD patients), patient understanding of the need to obtain care in these specialty institutions, and patient recognition of the overall need for lifelong care^8,13-15^.

While exceptional work has been done in 1) establishing recommendations for appropriate follow-up; 2) defining relationships between poor follow-up and increased morbidity, and 3) describing predictors for failing to maintain care^1,5,8,16,17^, many of these previous studies have focused on patients who had already re-engaged in care. Currently, no studies exist examining patients with CHD who are currently lost to follow-up. Thus, there is a gap in data for identifying chronic disease patients currently or “actively” missing from care and describing the barriers that keep them from re-engaging into the medical system.

As with many other chronic disease specialties, in pediatric cardiology we are not aware of who is actively missing from care, why they still have not returned for their follow-up, if they are being cared for elsewhere, or how they can be re-engaged in care during this period. The purpose of this study is to elucidate the answer to these questions for adolescents and young adults with CHD who remain missing from our cardiac care. We strive to define this population, understand their obstacles to access, and identify if a simple intervention could assist in re-engagement with us. Based on existing literature, we hypothesized that less severe CHD, longer intervals between follow-up, lower socioeconomic status, and being a racial/ethnic minority would result in higher rates of those ‘actively missing’ for transition-aged youth with CHD and that receiving a telephone prompt would assist in re-establishing cardiac care.

## METHODS

We conducted a retrospective single center cohort study with prospective phone interview follow-up for individuals we deemed to be actively missing from care. A review of outpatient electronic health records (EHR) from 2012 to 2019 of patients with CHD aged 15-21 years at the time of their most recent cardiology outpatient encounter at Texas Children’s Hospital (TCH) was conducted. Patients were excluded if they underwent a heart transplantation or ventricular assist device placement, lacked CHD, or scheduled follow-up within 1 year of our data collection. Identification of CHD diagnoses were determined by ICD 9/10 codes within the EHR, and diagnoses were stratified into disease complexity (simple, moderate, great) based on 2018 American Heart Association/American College of Cardiology adult congenital heart disease guidelines^17^. The primary outcome variable was being “actively missing” from cardiac care at TCH, which was defined as failure to return for outpatient cardiology follow-up ≥1 year beyond last requested clinic follow-up. Our primary predictor variables were patient sociodemographic data: sex, age (15-17 vs 18-21 years), race/ethnicity, primary language (English, Spanish, or other), CHD complexity, last cardiology provider (pediatric or adult congenital heart disease (ACHD)), insurance status (public, private, or other), and time to next requested follow-up (≤12 months vs >12 months).

The study underwent Internal Review Board approval before beginning data collection or conducting interviews. Interview scripts were created based on existing literature about why patients become lost to follow-up^8,11,14,18^. For patients identified as “actively missing” who we were able to reach and who consented to our study, a telephone interview was conducted to determine CHD knowledge, barriers to care, and current cardiac symptoms. Both young adult patients and/or their parents were permitted to complete the interview. Interviews were performed by four study co-authors (AA, AE, JM, JS) in the patient or parent’s preferred language with a licensed interpreter if conducted in a language other than English. The interview determined the current status of a patient’s cardiac care (in care vs not in care), and type of cardiologist (general adult, adult congenital, or pediatric cardiologist) managing their condition if they were in care. The interview then uses prompts to explore reasons patients missed their last appointment, barriers to scheduling a clinic visit, and reasons for not following up at our institution. Questions regarding recent or active cardiac symptoms (e.g. chest pain, difficulty breathing, palpitations, or recent syncope), medications, and level of confidence in describing their condition, were then asked. Lastly, a needs assessment was performed to determine most helpful interventions to assist with maintaining or re-engaging in care. At the completion of the interview, a simple intervention was performed: participants were offered information to reschedule follow-up for cardiac care by providing them the telephone number for the cardiology clinic scheduling line. Our intent was to see who, after being educated on ACHD services at TCH and the need for lifelong care, would 1) use this information to schedule a follow-up appointment and 2) attend a follow-up appointment within 4-6 months post scheduling.

## STATISTICAL ANALYSIS

Descriptive statistics were used to describe the actively missing population as well as those that participated in telephone interviews. Univariable and multivariable logistic regression models were used to determine significant associations between sociodemographic variables and the outcome variable of those who were presently in care vs those actively missing from care. Variables felt to be potential factors associated with the outcome were chosen a priori based on existing literature and clinical observations. Covariates in the univariate model with a p value <0.1 were entered into the multivariate model. A p value <0.05 was considered statistically significant. SPSS version 26 was used for statistical analysis.

## RESULTS

### Descriptive, univariate, and multivariate analyses

#### Demographics of those “actively missing” from care

From 2012-2019, a total of 1053 patients with CHD were seen between the ages of 15-21 years with requested return follow-up and 67% maintained care (Table 1). Within the primary cohort, 349 patients were identified to be actively missing (33%). The median age of those actively missing was 17 years, and 203 patients (58%) were male. Forty-six percent (n=159) of patients were Non-Hispanic White, 33% (n=113) were Hispanic, and 9% (n=33) were non-Hispanic Black. Fifteen percent identified their primary language to be Spanish. Seventy-eight percent (n=278) of patients were still under the care of a pediatric cardiologist when last seen, and 62% (n=217) of these patients noted having private insurance. We attempted telephone contact with all actively missing patients to conduct our interview. A total of 125 actively missing patients and/or their parents (36%) were able to be reached and willing to complete the interview.

**Table 1.**
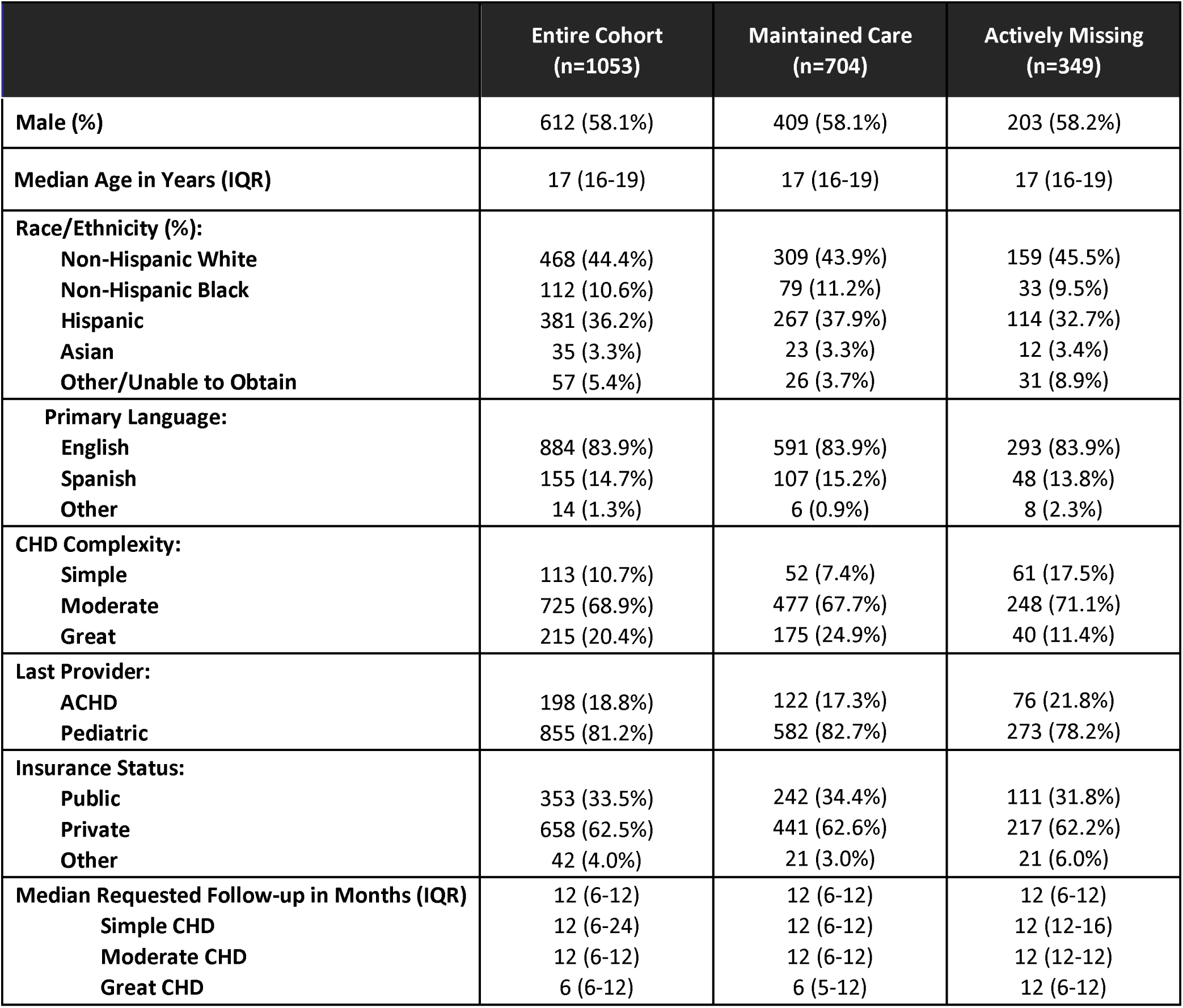
Patient demographics and follow-up characteristics

#### Associations between sociodemographic factors and ‘actively missing’ from care

On univariate analysis, age, race/ethnicity, CHD complexity, type of provider, insurance status, and requested timing of follow-up were all statistically significantly variables associated with being actively missing from cardiac care (Table 2). On multivariable analysis, age, CHD complexity, type of provider, and timing of follow-up remained statistically significant. Older patients (18-21 years) at the time of their last visit were over four times more likely to be actively missing from care. CHD complexity was associated with being actively missing from care: patients with simple forms of CHD were greater than 2.5 times more likely to be actively missing when compared to great complexity patients. Time to next requested follow-up of >12 months resulted in patients being 1.5 times more likely to be actively missing when compared to those requested to follow-up within 12 months. Lastly, if the most recent cardiologist was an ACHD provider, patients were more likely to be actively missing. Interestingly, sex, race/ethnicity, primary language, and insurance status were not associated with being actively missing from cardiac care.

**Table 2.** Univariable and multivariable analysis of factors associated with actively missing patients

|  | Maintained Care<br>(n=704) | Actively Missing<br>(n=349) | Univariate |  | Multivariate |  |
| --- | --- | --- | --- | --- | --- | --- |
|  |  |  | OR (CI) | p value | OR (CI) | p value |
| **Sex:** |  |  |  |  |  |  |
| Male | 409 (58.1%) | 203 (58.2%) | 1.00 (0.73-1.30) | 0.98 | - | - |
| Female | 295 (41.9%) | 146 (41.8%) | ref | - | - | - |
| **Age:** |  |  |  |  |  |  |
| 15-17 years | 502 (71.3%) | 116 (33.2%) | ref | - | ref | - |
| 18-21 years | 202 (28.7%) | 233 (66.8%) | 4.99 (3.78-6.58) | <0.001 | **4.37 (3.26-5.85)** | **<0.001\*** |
| **Race/Ethnicity:** |  |  |  | **0.009** |  | 0.433 |
| Non-Hispanic White | 309 (43.9%) | 159 (45.5%) | ref | - | ref | - |
| Non-Hispanic Black | 79 (11.2%) | 33 (9.5%) | 0.81 (0.52-1.27) | - | 1.15 (0.62-2.11) | - |
| Hispanic | 267 (37.9%) | 114 (32.7%) | 0.83 (0.62-1.11) | - | 1.30 (0.63-2.69) | - |
| Asian | 23 (3.3%) | 12 (3.4%) | 1.01 (0.49-2.09) | - | 1.55 (0.84-2.87) | - |
| Other/Unable to Obtain | 26 (3.7%) | 31 (8.9%) | 2.32 (1.33-4.04) | - | 1.18 (0.46-3.05) | - |
| **Primary Language:** |  |  |  | 0.158 |  |  |
| English | 591 (83.9%) | 293 (83.9%) | ref | - | - | - |
| Spanish | 107 (15.2%) | 48 (13.8%) | 0.91 (0.63-1.31) | - | - | - |
| Other | 6 (0.9%) | 8 (2.3%) | 2.69 (0.93-7.82) | - | - | - |
| **CHD Complexity:** |  |  |  | **<0.001** |  | **0.002\*** |
| Simple | 52 (7.4%) | 61 (17.5%) | 5.13 (3.09-8.50) | - | 2.70 (1.55-4.71) | - |
| Moderate | 477 (67.7%) | 248 (71.1%) | 2.28 (1.56-3.31) | - | 1.59 (1.06-2.40) | - |
| Great | 175 (24.9%) | 40 (11.4%) | ref | - | ref | - |
| **Last Provider:** |  |  |  |  |  |  |
| ACHD | 122 (17.3%) | 76 (21.8%) | 1.33 (0.96-1.83) | **0.083** | 1.53 (1.07-2.19) | **0.020\*** |
| Pediatric | 582 (82.7%) | 273 (78.2%) | ref | - | ref | - |
| **Insurance Status:** |  |  |  | **0.060** |  | 0.166 |
| Public | 242 (34.4%) | 111 (31.8%) | ref | - | ref | - |
| Private | 441 (62.6%) | 217 (62.2%) | 1.07 (0.81-1.42) | - | 0.81 (0.58-1.13) | - |
| Other | 21 (3.0%) | 21 (6.0%) | 2.18 (1.14-4.16) | - | 1.43 (0.70-2.91) | - |
| **Requested Follow-up:** |  |  |  |  |  |  |
| ≤12 months | 644 (91.5%) | 275 (78.8%) | ref | - | ref | - |
| >12 months | 60 (8.5%) | 74 (21.2%) | 1.75 (1.25-2.44) | **0.001** | 1.49 (1.03-2.16) | **0.036\*** |

### Telephone interviews with those “actively missing” from care

#### Cardiology care

Among the n=125 interview responders, 61% (n=76) were currently not being seen by any cardiologist. Of the 39% (n=49) actively being cared for a cardiologist outside our hospital, nearly half (43%; n=21) were followed by an adult cardiologist, while 24% (n=12) were followed by an ACHD provider, and 33% (n=16) by a pediatric cardiologist. When asked about cardiac symptoms, nearly a quarter of patients (21%; n=26) reported recently feeling one of the described cardiac symptoms (chest pain, syncope, shortness of breath, or palpitations), with chest pain being most common (42%, n=11). Despite these former patients reportedly having recent cardiac symptoms, 21 (81%) of them were not receiving any cardiac care at the time of interview. Twenty-five patients (19%) reported being on cardiac medications with 84% (n=21) of those on medications reporting be able to name them. Most responders felt comfortable explaining their condition to someone else, with 58% (n=72) reporting greater than 3 on a 1-5 Likert scale, and an overall mean score of 3.6 (iQR: 3-5).

#### Reasons for missing follow-up appointments

Among those not currently being cared for by a cardiologist, the most common reasons cited for missing appointments were: lack or change of insurance (24%, n=18), inability to afford clinic visit payments (20%, n=15), or lack of symptoms (14%, n=11) (Figure 1). When asked regarding ease of rescheduling cardiology appointments, 30% (n=38) noted scheduling obstacles that deterred their follow-up. Among all the responders, specific barriers preventing them from being seen at our institution included: 1) having insurance that was not accepted at TCH (35%, n=44); 2) moving outside the local area (23%, n=29); and 3) being unaware of ACHD services at our hospital (20%, n=25).

**Figure 1.**
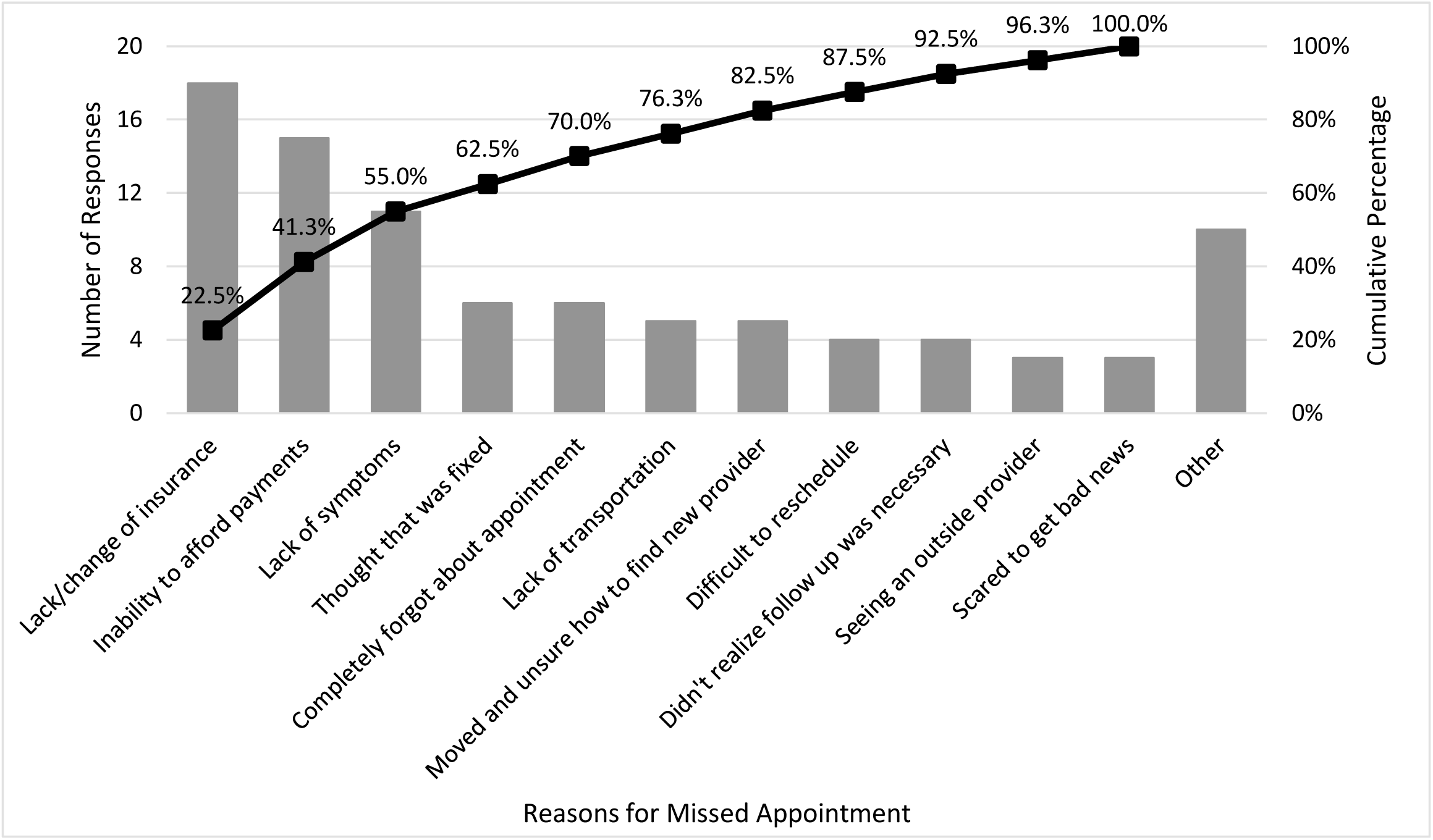
Pareto chart of interview responses of reasons for missing cardiology appointments.

### Simple intervention

Interview responders were given information about the need for lifelong CHD care and offered the scheduling line for the TCH cardiology clinic in order to re-engage in care. Of those offered the telephone number, 55% (n=69) accepted the information. Four to six months after the interview, the EHR was reviewed to determine which of those patients had scheduled cardiology clinic appointments. Only 7 responders (10%) had actually scheduled an appointment with only 2 patients (3%) successfully re-engaged in care at the TCH cardiology outpatient clinic.

## DISCUSSION

Obstacles that prevent cardiology patients from maintaining appropriate cardiac care are diverse and challenging to address, particularly during the transition period when transferring from pediatric to adult cardiac providers. Our data on actively missing patients reveals the highest risk group are patients in their late teens to early twenties with simple to moderate CHD, whose next cardiology follow-up is >12 months from their last visit, and who have recently transferred to an adult cardiac provider.

Inconsistency of care in medically complex patients across different sexes, races/ethnicities, and insurance statuses has been previously published^11,14,19^. In our study, race/ethnicity, primary language, and insurance type were not associated with decreased outpatient follow-up. This may be because other sociodemographic factors were more impactful on multivariable analysis (age, provider type, disease severity, etc), and it is difficult to know frequency of parental insurance changes due to job transitions, losses or “aging out of the public insurance system”, but data has shown that as a patient with CHD ages, there are still likely to be disparities in insurance coverage, particularly for transition aged minority patients with CHD^13^. Simpler CHD is a patient characteristic that has been previously associated with either gaps in care or loss to follow-up^7,11,14^. However, previous studies demonstrate variability in follow-up rates across cardiac centers, especially those with lower outpatient volumes^1,8,11,14^. For adolescents and young adults with CHD, data has shown that not only has hospital admission volume increased 48.7% between 2009 to 2013, but that inpatient admissions have risen more sharply for those with non-severe CHD compared to severe CHD^20^. It is critical that patients understand simple lesions still require lifelong follow-up, and that understanding/strategizing ways to maintain insurance coverage while in pediatric cardiology care are key to reducing gaps in transition aged and adult care.

Telephone interview responses demonstrate patients/parents felt confident in CHD knowledge: the ability to describe their CHD and list medications. Yet having a lack of symptoms and/or awareness of lifelong care as motives for absence does suggest a disconnect in comprehending the potential prognosis of their disease. Despite being one of only two high volume ACHD centers (at time of study period ending in 2019) in the second most populous state in the United States, institutional limitations in accepted insurance plans and lack of public awareness of accredited centers for ACHD care has constrained our ability to care for our transition-aged and ACHD patients^21^. If these hardships remain unaddressed, the ability to care for our patients will continue to be hindered and inadequate.

It is evident institutional practices regarding ease of scheduling, frequency of follow-up, and understanding needs for specific ACHD care even at pediatric institutions contribute to patients being lost from cardiac care based on the responses collected in our study. More broadly, general obstacles to both keeping an appointment and rescheduling were further fraught with difficulty navigating the medical system and lack of awareness to local subspecialist resources. Considering how few patients were able to re-enter care from the provided scheduling line, it seems a more streamlined and “navigated” access structure is necessary for transition-aged patients. Methods for re-capturing currently missing patients as well as actively assisting them in re-engaging in appropriate clinical care are key components to a more suitable strategy. Furthermore, a transition education program assisting in patient empowerment and understanding the need for lifelong care, creating a transfer summary and maintenance of care plan, helping patients navigate a clear and straightforward method for scheduling and rescheduling appointments, and financial counseling to assist in insurance maintenance are critical to improving continuity of care and patient outcomes.

The ability to schedule online and text message reminders that directly provide options for confirmation or rescheduling was preferred by our responders and can facilitate future organization of follow-up care. Another way to facilitate care could be patient portals to ease scheduling or streamline financial counseling. Working through transition programs to help patients learn how to best maintain health insurance, providing options for financial assistance, and/or guidance on mitigation of healthcare costs would be of great benefit based on reported barriers. Collaboration with transition teams, social work services, patient and family advocacy groups, hospital administration, as well as federal, state, and local care insurance coverage policies may assist in accomplishing the goal of maintaining care. Evaluating federal and state policies for maintenance of insurance and safety net options for those without insurance are critical systemic necessary changes as well. Transition education and skill building are imperative for patients to understand the need for future care and what appropriate ACHD care looks like. Pediatric cardiologists should be individually aware of those potentially at highest risk to drop out of care and devise a maintenance of care and transfer of care plan. Patients cared for by pediatric cardiologists should have identified ACHD providers prior to transfer to adult care and should be provided available resources within the AHCD community, such as the adult congenital heart association (ACHA) website (https://www.achaheart.org/)^22^. For those cardiologists who do not have formal transition programs, there is a clear need for educating patients with the assistance of online or mobile-based transition programs that discuss obstacles and strategize access to ACHD care, insurance concerns, patient self-empowerment, and the importance of follow-up.

## LIMITATIONS

There a number of limitations associated with this study. The retrospective data collection to identify our study group is reliant on accurate ICD coding and appropriate selection of requested follow-ups. Additionally, this is a cross-sectional study that represents an isolated point in time and therefore is not designed to represent every patient’s individual gaps in care or their duration of absence. The telephone interview data is limited to only those able to be contacted and willing to participate. Sixty-seven percent of those actively missing patients could not be represented in the interview data, which speaks to the need to capture important data, barriers, and social determinants of health for patients before or immediately once they fall out of care. Additionally, interview practices could potentially introduce bias, but uniform scripts and limited numbers of interviewers were utilized. The current Coronavirus Disease 2019 (COVID-19) pandemic could have also contributed to the lack of follow-ups identified after the scheduling information was provided to interview participants, although the majority of patient follow up 4-6 most post interview preceded the pandemic, and this was a rarely cited reason in our interviews. Finally, each institution’s local challenges, patient population, ACHD physician and ACHD accredited center access, local and state health policies, and community support structure is different, which may limit the generalizability of these results.

## CONCLUSION

Adolescents and young adults with CHD live complicated lives affecting their ability to maintain cardiac care, but patient demographics alone (e.g. sex, race/ethnicity, language, or insurance status) fail to encompass these challenges. Scheduling practices, insurance acceptance at CHD facilities, public awareness of ACHD centers, length of time between visits, and providers are all reported barriers determined by healthcare. Transition programs could assist in streamlining some aspects of this education, including identifying an ACHD specialist prior to transfer of care, as could scheduling through a patient portal or online/telephone service for established CHD patients. The medical team and patients/families share a responsibility to address obstacles to access, and merely providing clinic contact information is inadequate to reintroduce them to appropriate cardiology follow-up.

## Data Availability

The data obtained for this study was obtained internally. No external datasets were used.

## Abbreviations

CHD: Congenital Heart Disease
ACHD: Adult Congenital Heart Disease
EHR: Electronic Health Record
TCH: Texas Children’s Hospital
ACHA: Adult Congenital Heart Association
COVID-19: Coronavirus Disease 2019

